# Assessment of MRI Geometric Distortion for Radiotherapy Planning

**DOI:** 10.64898/2026.07.30.26359350

**Authors:** Mariia Odnovol, Ekaterina N. Lykova

## Abstract

**Background:** MRI is widely used in radiotherapy planning due to its high soft-tissue contrast, but geometric distortions can compromise target localization accuracy.

**Methods:** Two phantoms – a commercial anthropomorphic STEEV phantom and a custom-made phantom fabricated from ABS plastic – were scanned on MRI and CT. Distortion was measured via linear distances between periodic structures in a DICOM viewer, with CT as the reference. Dose modelling was performed to assess the effect of target displacement on D99% coverage.

**Results:** STEEV phantom showed ≤0.95 mm maximum distortion (≤0.2 mm within 30 mm from isocenter). The custom phantom showed 0.30 ± 0.05 mm in the central zone (within 50 mm) and up to 3.1 mm at the periphery (102 mm from center). For a 0.1 cm^3^ target, a 1 mm shift reduced D99% from 95% to 82%; for a 5.2 cm^3^ target the reduction was negligible.

**Conclusions:** Distortion increases with distance from isocenter and is clinically significant for small targets (<1.6 cm^3^). The custom ABS phantom offers a cost-effective tool for routine quality assurance. The proposed method is accessible for resource-limited settings.

## Introduction

Magnetic resonance imaging is widely employed in radiotherapy owing to its high soft-tissue contrast, which enables more precise visualization of tumor targets and critical structures. However, MRI possesses a significant limitation – the presence of geometric distortions, attributable to inhomogeneity of the static magnetic field, gradient field nonlinearity, chemical shift, and other physical effects [1].

For radiotherapy, particularly in stereotactic radiosurgery where high spatial precision is essential, even minor distortions may prove critical. Failure to account for distortion can result in systematic errors in target localization, thereby reducing treatment efficacy and increasing the risk of complications. Computed tomography, in contrast to MRI, exhibits minimal geometric distortions due to the rigid geometry of the X-ray tube and detectors; consequently, CT images may serve as a reference standard for assessing MRI distortion [2, 3].

**Aim of the study** – to measure distortion on MR images using two phantoms and to evaluate its impact on radiotherapy planning.

### Research objectives

1. Review of the scientific literature on MRI distortion.
2. Selection of a method for measuring distortions in head scanning.
3. Development and evaluation of a custom-made phantom from ABS plastic.
4. Scanning of the commercial STEEV phantom and the custom-made phantom on MRI and CT scanners.
5. Measurements of the acquired images.
6. Comparative analysis of MRI and CT images.
7. Assessment of distortion relative to reference CT data.
8. Quantitative evaluation of the impact of distortion on radiotherapy treatment plans.

## Materials and methods

Two phantoms were employed in the study.

The first was a commercial anthropomorphic STEEV phantom (CIRS Inc., USA), simulating human body structures. A cubic insert measuring 63*63*93 mm^3^ with periodic internal structures was utilized for measurements, enabling distortion assessment in the central region of the scanner.

The second phantom was designed and fabricated by the author from ABS plastic (acrylonitrile butadiene styrene). Its overall dimensions are 143*192*95 mm^3^, with periodic internal structures (rows of cubes) for tracking spatial distortions. The selection of ABS plastic was motivated by its financial availability, ease of machining, and high hydrogen proton content, which provides an adequate signal on MR images. This phantom is intended for distortion assessment across the entire field of view, including peripheral regions.

The combination of two phantoms of differing dimensions enables comprehensive distortion assessment: STEEV provides local measurements at the center, while the custom-made phantom provides global measurements across the entire field of view.

**Fig. 1.**
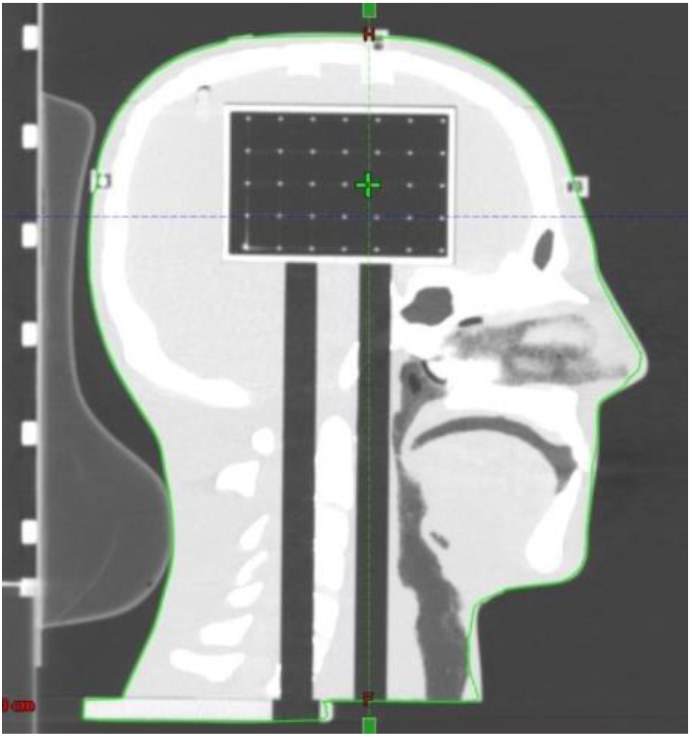
“Anthropomorphic STEEV phantom (CT image)”

**Fig. 2.**
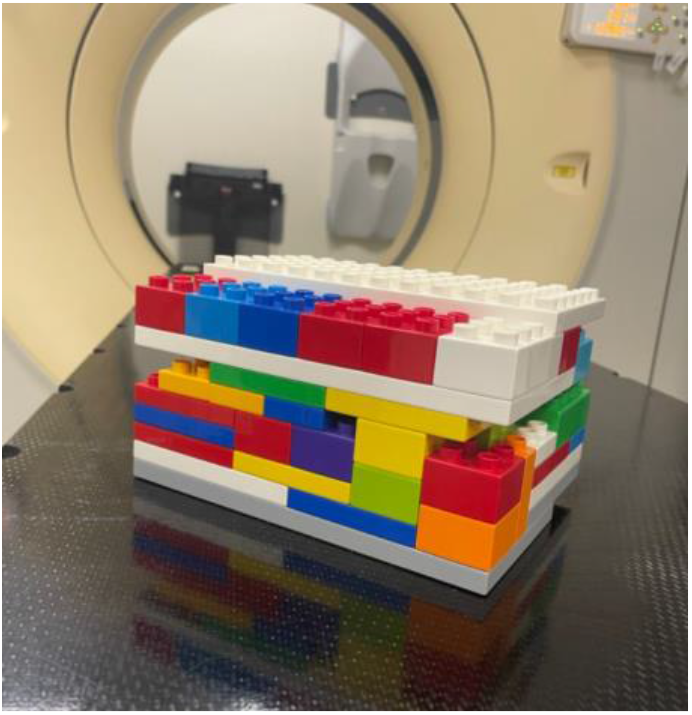
“Custom-made phantom (ABS-plastic); scanning machine: MRI Siemens MAGNETOM Aera, CT Phillips Brilliance iCT; European Medical center (Moscow, RU);”

CT images were utilized as the reference standard due to the absence of significant geometric distortions, attributable to the rigid geometry of the system. This study was conducted using phantoms only; no human subjects or patient data were involved.

## Image processing

Image processing was conducted in two stages: primary delineation in a DICOM viewer and subsequent statistical analysis in Microsoft Excel.

The processing sequence included:

1. Structure identification — periodic rows of structures (cubes) were visually identified on axial and sagittal MRI and CT slices for both phantoms.
2. Linear measurement – distances between corresponding control points (edges of structures) along the OX and OZ axes for each row were measured on both image types using the calibrated linear measurement tool integrated into the DICOM viewer. Measurements were performed for all available rows across the entire field of view.
3. Absolute distortion calculation – for each measured distance, the absolute difference between the values obtained on MRI and the reference CT was computed: Δ = |L_MRT - L_CT|. Calculations were performed in Microsoft Excel.
4. Measurement error estimation — the accuracy of manual delineation in the DICOM viewer was taken as half the voxel size, corresponding to ± 0.05 mm. This value was adopted as the systematic measurement error.
5. Density profile analysis — to verify the identity of the measured structures and assess spatial correspondence, one-dimensional density profiles were constructed for both imaging modalities. For CT, the Hounsfield scale (HU) was used; for MRI, signal intensity was used. The profiles were plotted along the same spatial coordinates for corresponding CT and MRI slices. Density profiles demonstrated coincidence of peaks on CT and MRI, confirming correct identification of structures by both methods. For the STEEV phantom, the peaks on CT and MRI density profiles coincided, confirming accurate spatial correspondence in the central region. For the custom-made ABS phantom, peaks on CT and MRI profiles were found to be mismatched at peripheral locations, indicating systematic geometric distortion.
6. Graph plotting — based on the obtained linear measurement data, graphs of distortion magnitude (Δ) versus distance from the scanner isocenter along the OX and OZ axes were plotted in Excel for both phantoms. These graphs demonstrated that distortion was minimal in the central zone (within 30–50 mm from isocenter) and increased progressively toward the periphery.
7. Dose modeling — based on the obtained distortion values, a simulation of the impact of geometric target shift on the D99% parameter (dose covering 99% of the PTV volume) was performed using an analytical model of target center displacement.

**Fig. 3.**
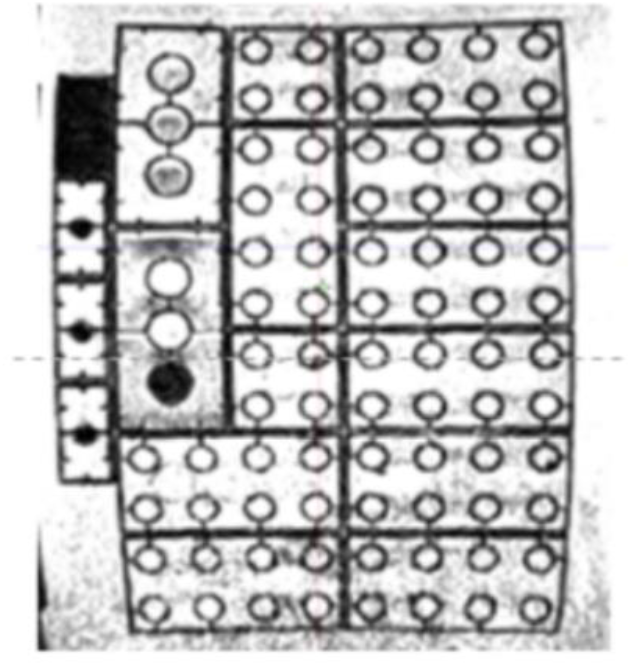
“Axial MRI slice acquired on a Siemens MAGNETOM Aera system at 1.5 T Scanning mode: T1-weighted imaging”

**Fig. 4.**
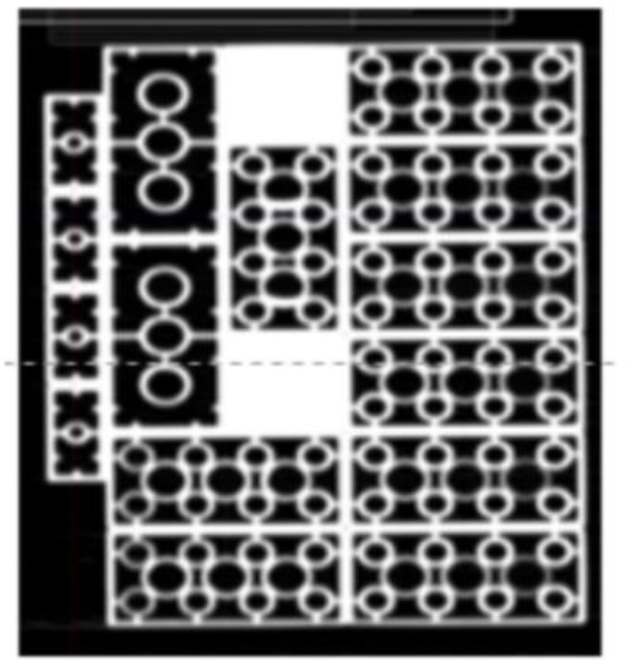
“Axial CT slice of the custom-made ABS phantom acquired on a Philips Brilliance iCT system.

**Fig. 5,6.**
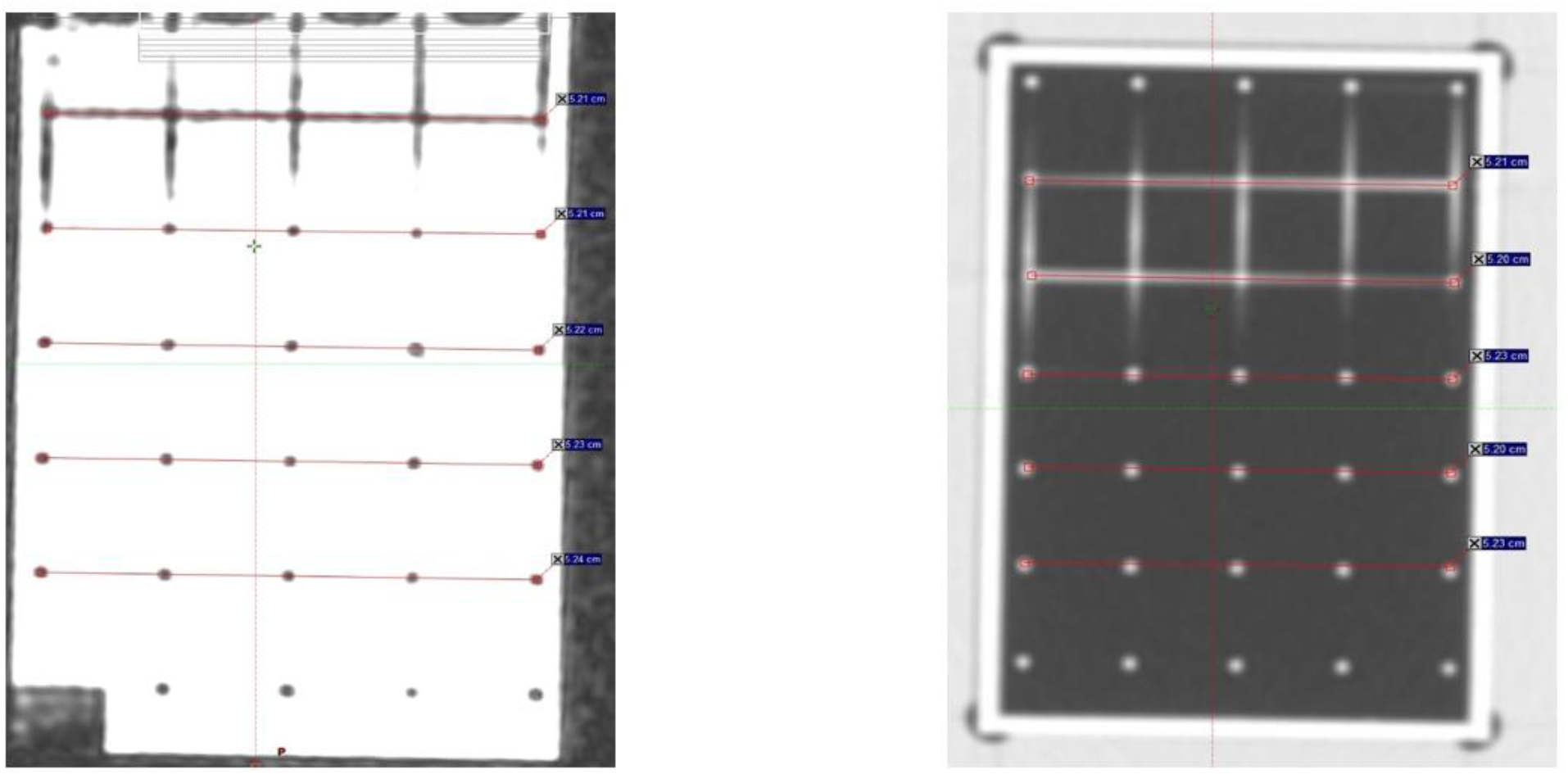
“Linear measurements via DICOM viewer: MR-image on the left, CT-image on the right”

**Fig. 7.**
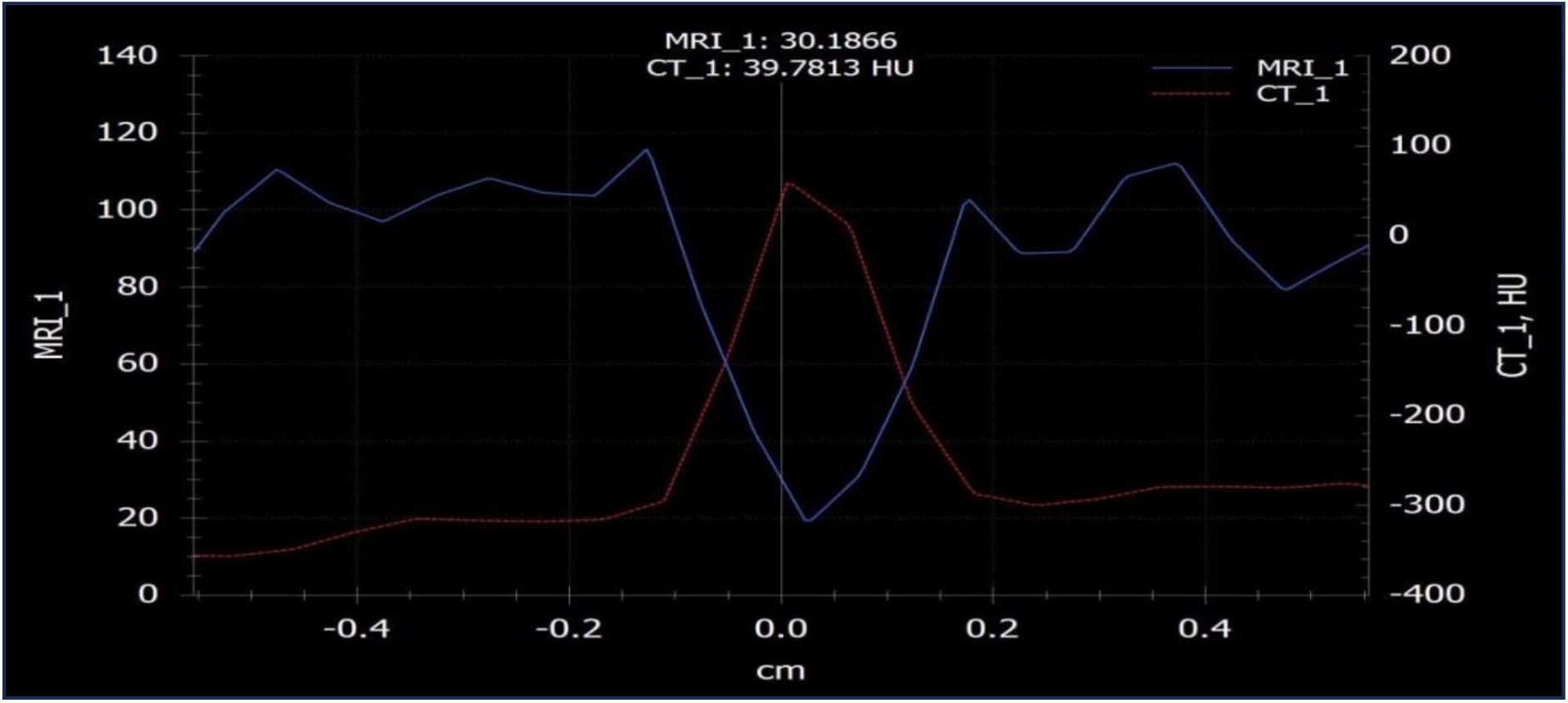
“Graph of the dependence of tissue density on the coordinate on MRI and CT (STEEV phantom)”

**Fig. 8.**
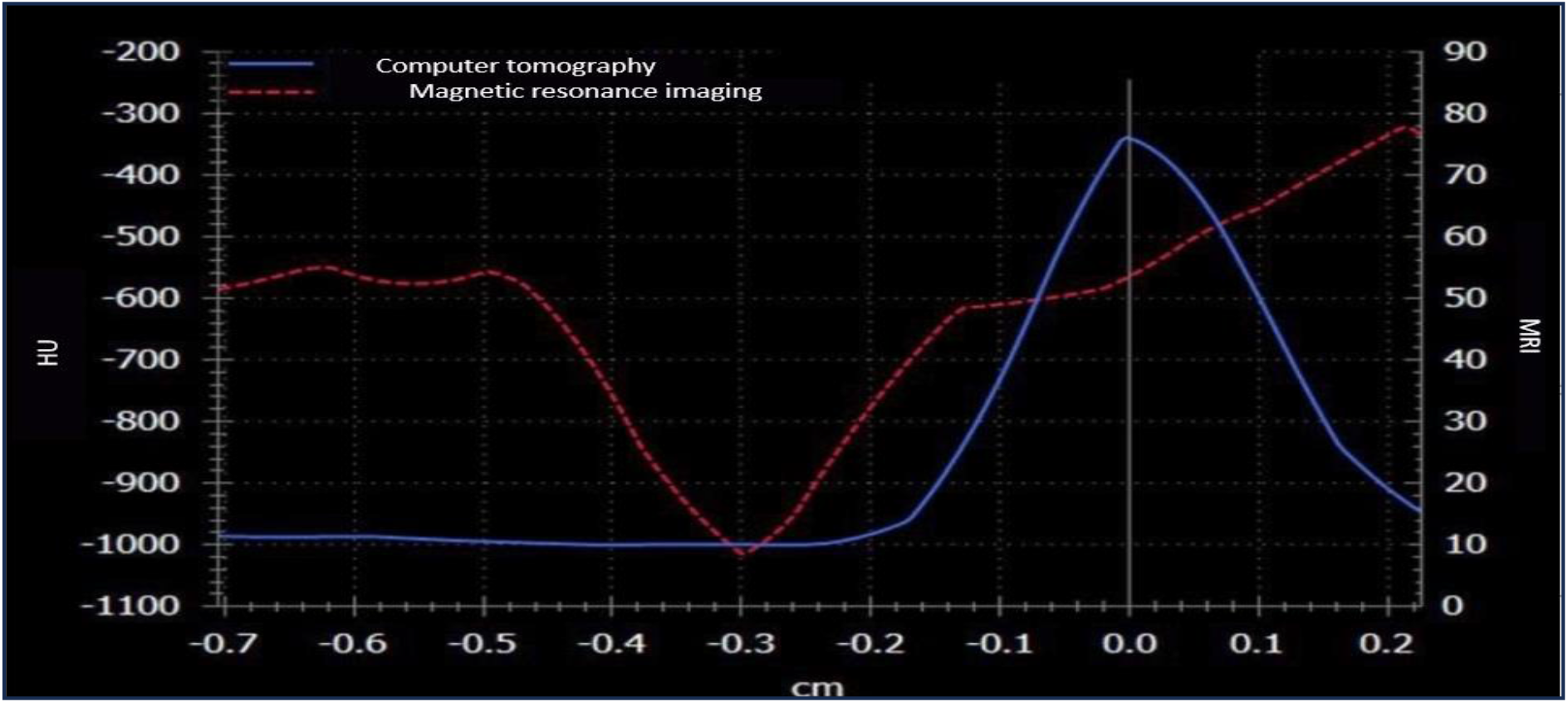
“Graph of the dependence of tissue density on the coordinate on MRI and CT (ABS-plastic phantom)”

**Fig. 9.**
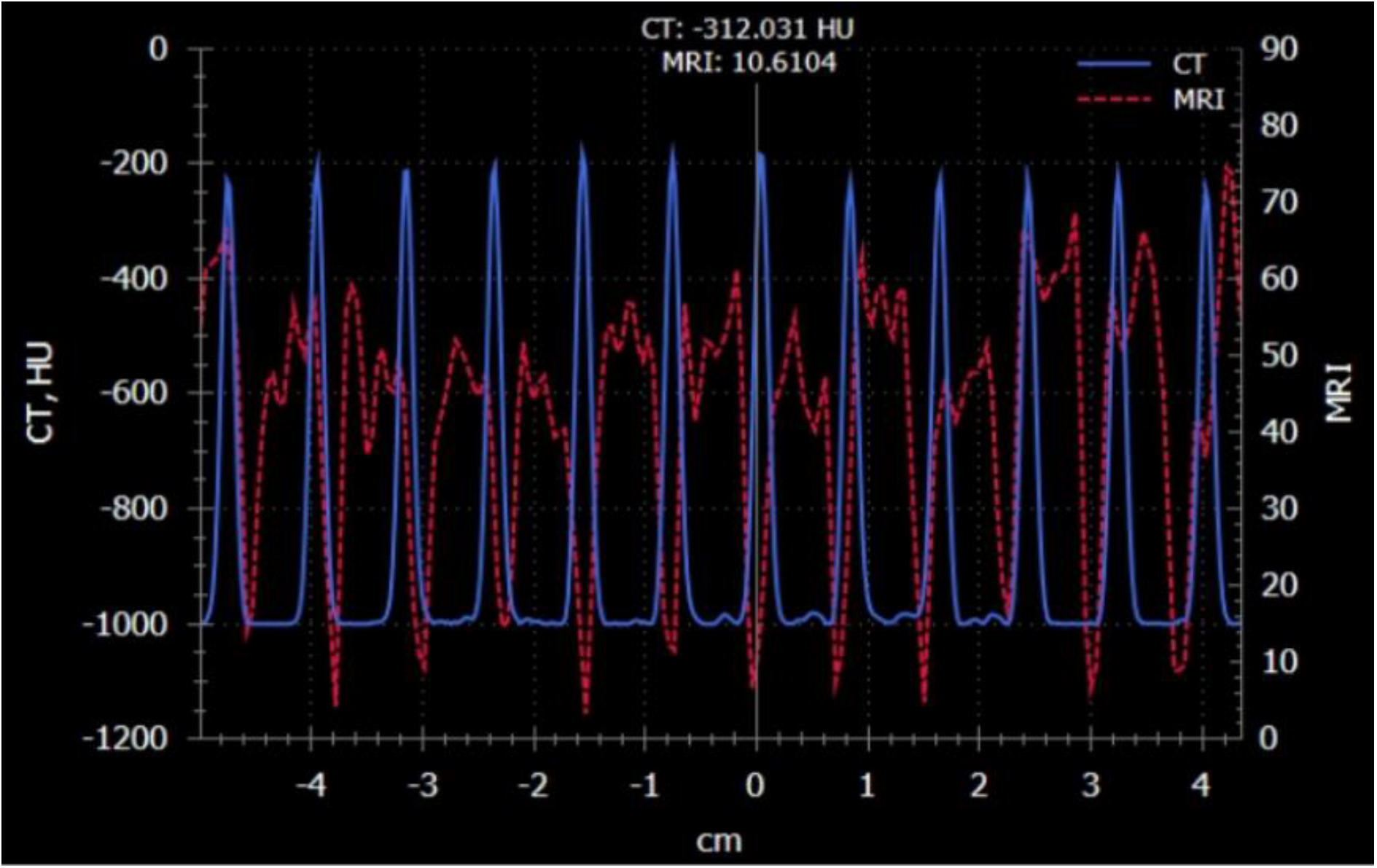
“Graph of the dependence of tissue density on the coordinate on MRI and CT (ABS-plastic phantom); more coordinates are used for detection”

## Results

### 1. Custom-Made ABS Phantom

For the custom-made phantom, a clear spatial dependence of distortion was revealed, confirming the results obtained with STEEV but with greater amplitude at the periphery. In the central zone within 50 mm from isocenter, the mean distortion was 0.30 ± 0.05mm. At the periphery (beyond 50 mm from isocenter), deviations reached 1.50 ± 0.05 mm or more. The maximum recorded values at the periphery were: in the axial plane, distortion reached 3.1 mm at a distance of 102 mm from the center; in the sagittal plane – 1.8 mm.

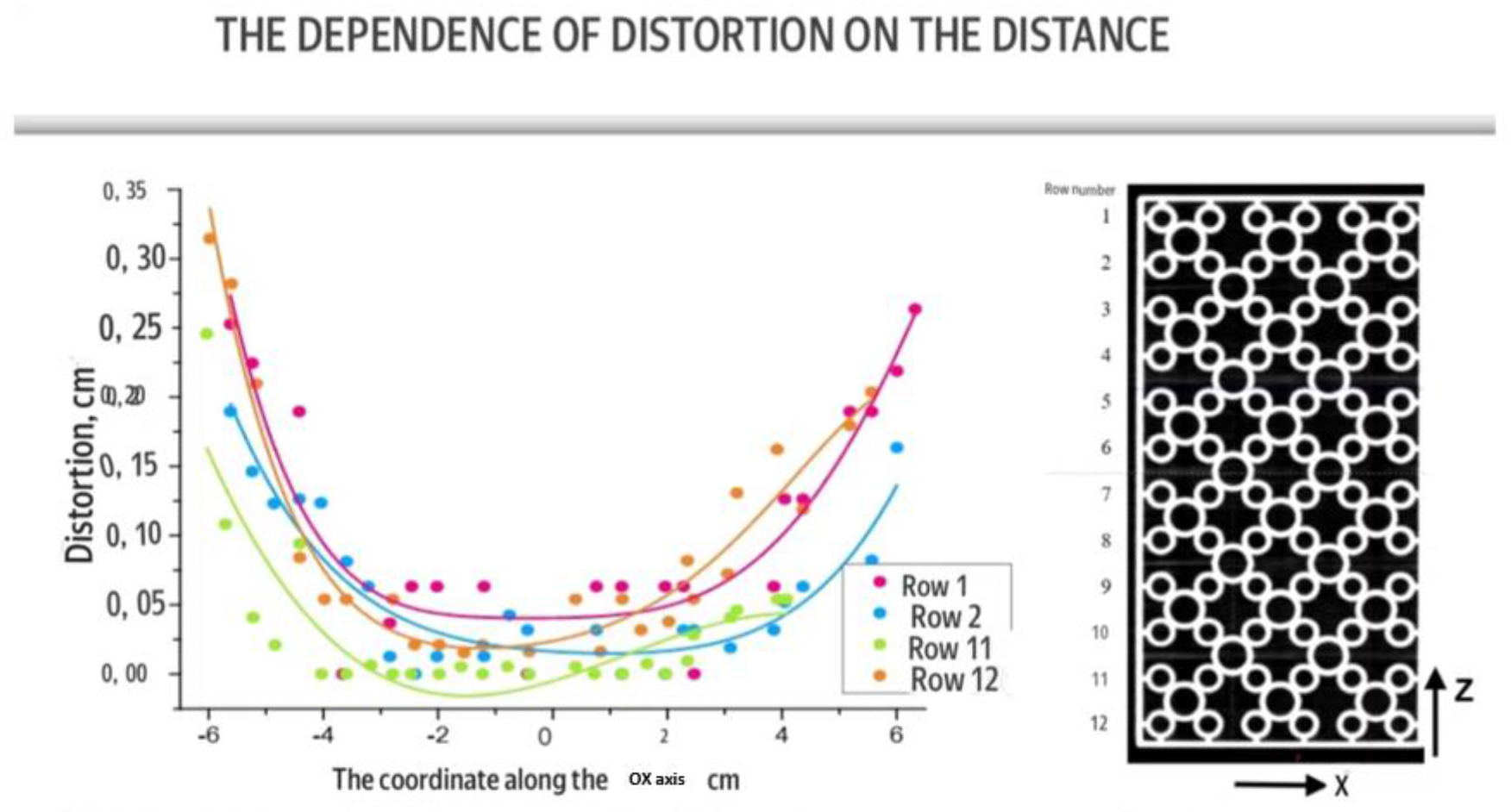
**Fig. 10.** Graph of the dependence of the distortion of the MRI image along ox_axis_ on the distance to the central OZ axis in the axial slice **Fig. 11.** Arrangement of rows of phantom structures

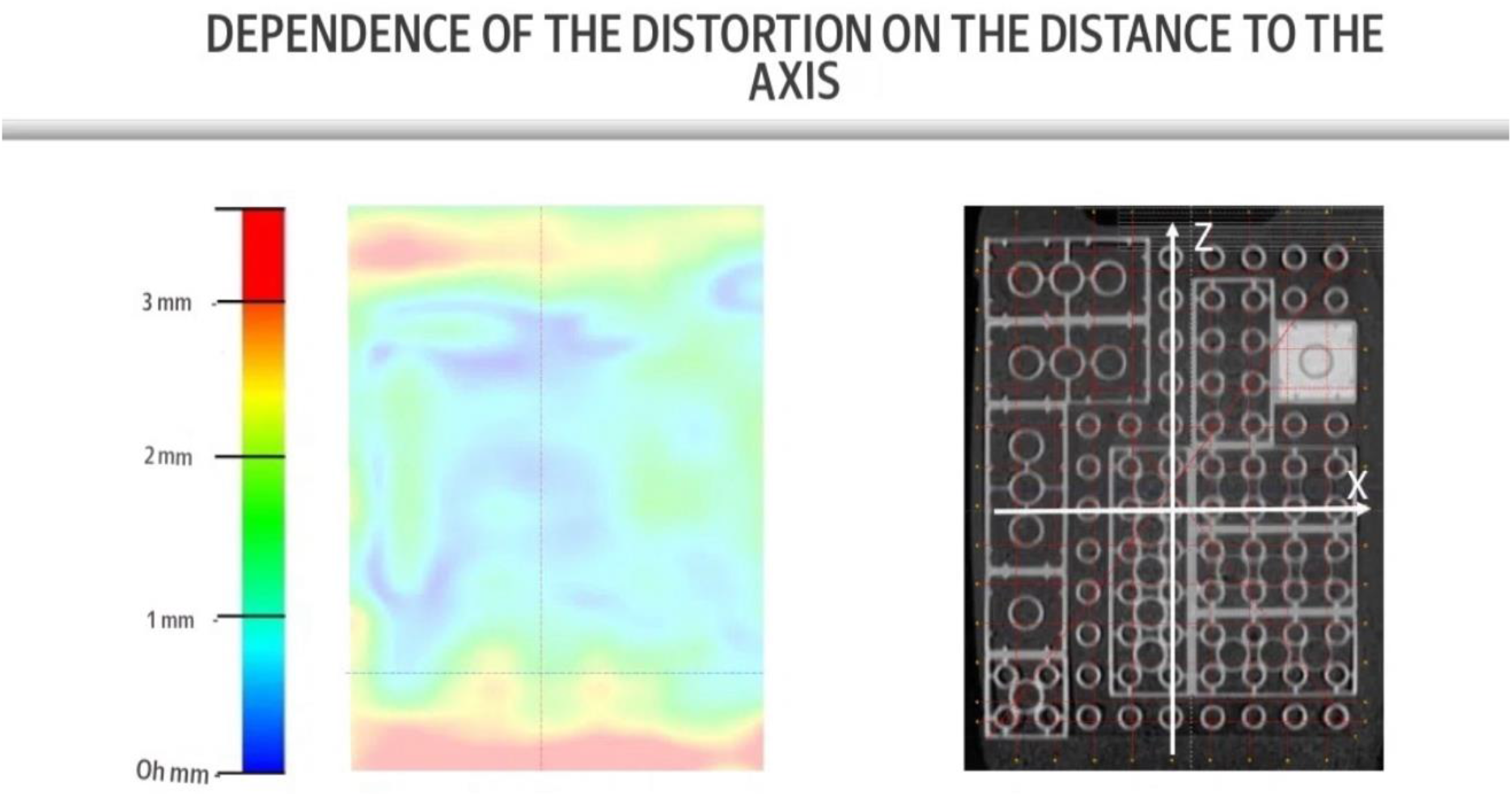
**Fig. 12.** Distortion map on the axial slice of the phantom **Fig. 13.** Axial slice of the phantom image

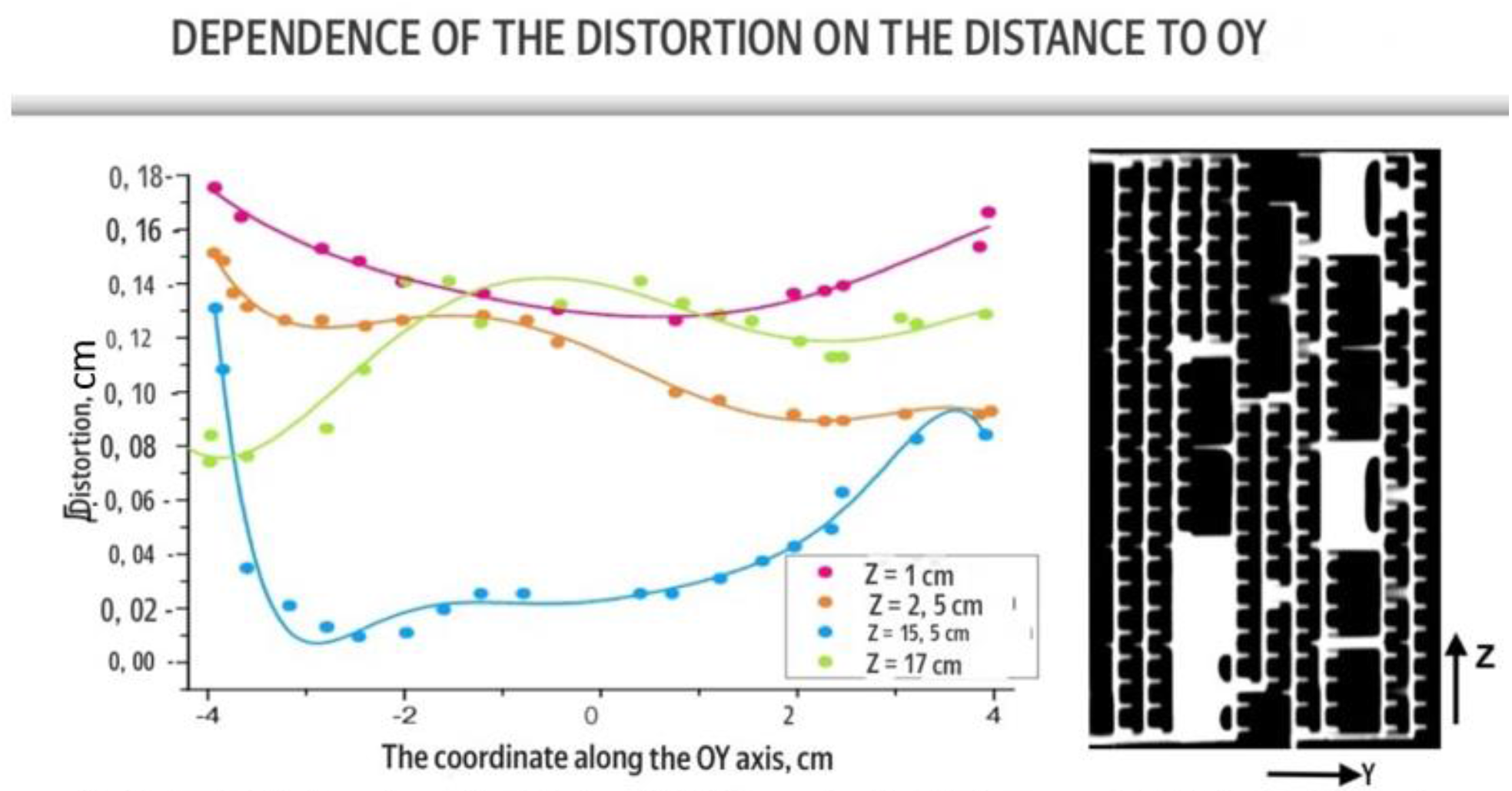
**Fig. 14.** Graph of the dependence of the distortion of the MRI image along the OZ axis on the distance to the central OY axis in the sagittal section. **Fig. 15.** The location of the axes in the image

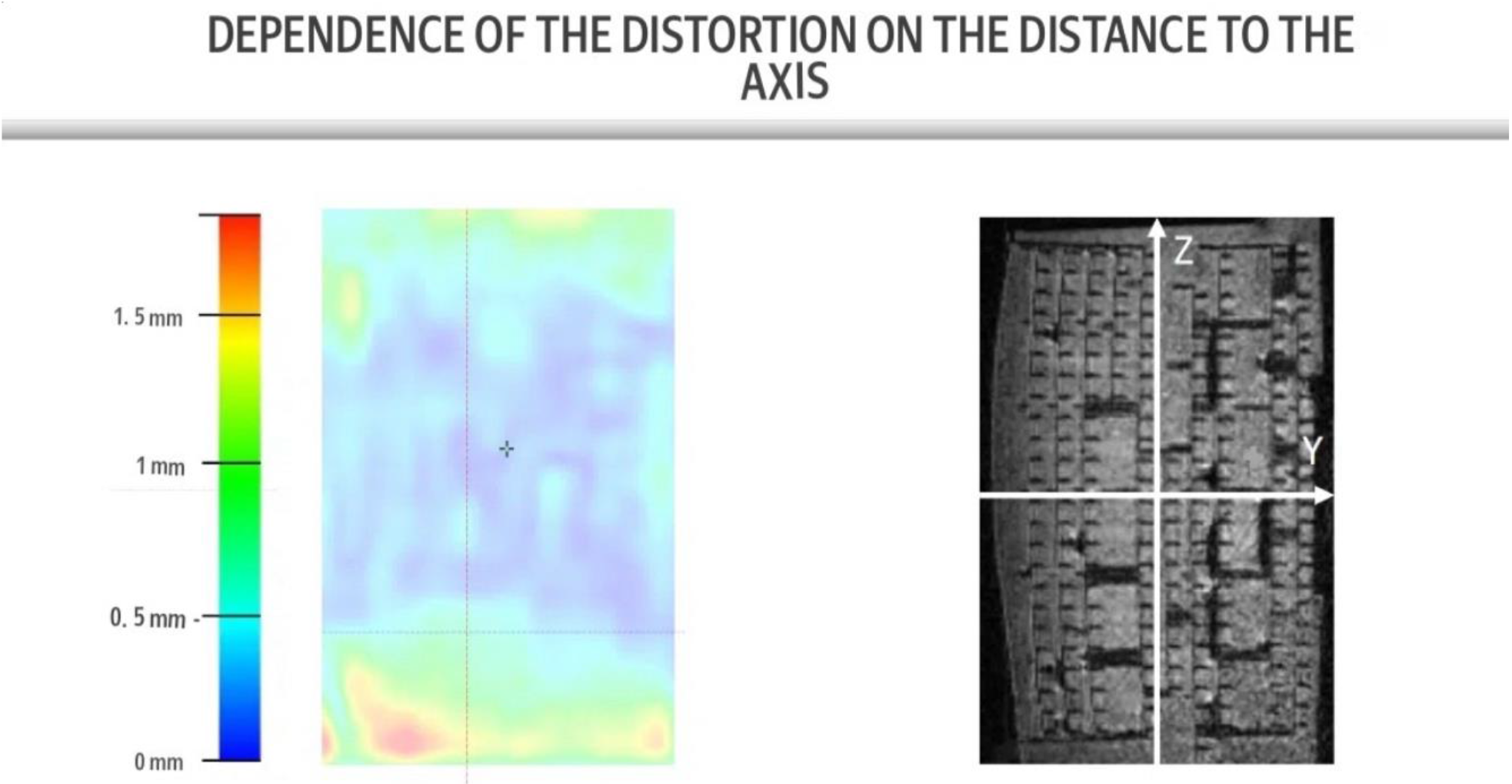
**Fig. 16.** Distortion map on the sagittal section of the phantom **Fig. 17.** Sagittal slice of the phantom image

### 2. Anthropomorphic commercial phantom STEEV

Linear measurements of the periodic structures of the STEEV phantom revealed that in the central region of the scanner, discrepancies between MRI and CT are minimal. As distance from the isocenter increases, distortion growth was observed. The maximum absolute deviation of linear dimensions across all measured rows did not exceed 0.95 mm. In the central zone (within 30 mm from isocenter), distortions did not exceed 0.2 mm, confirming the high linearity of the gradient system near the magnet center.

**Fig. 18.**
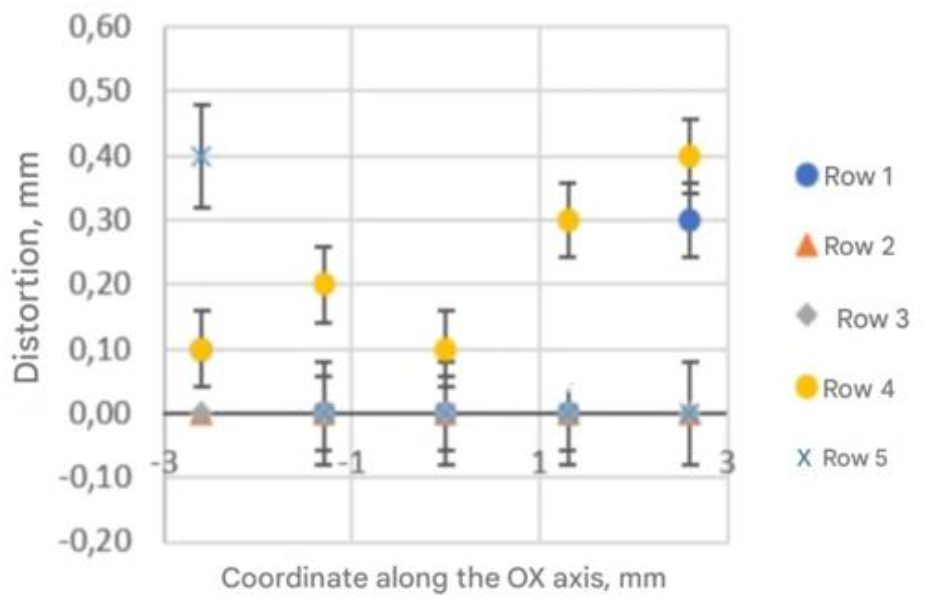
“Graph: dependence of distortion on coordinate (STEEV)”;

**Fig. 19.**
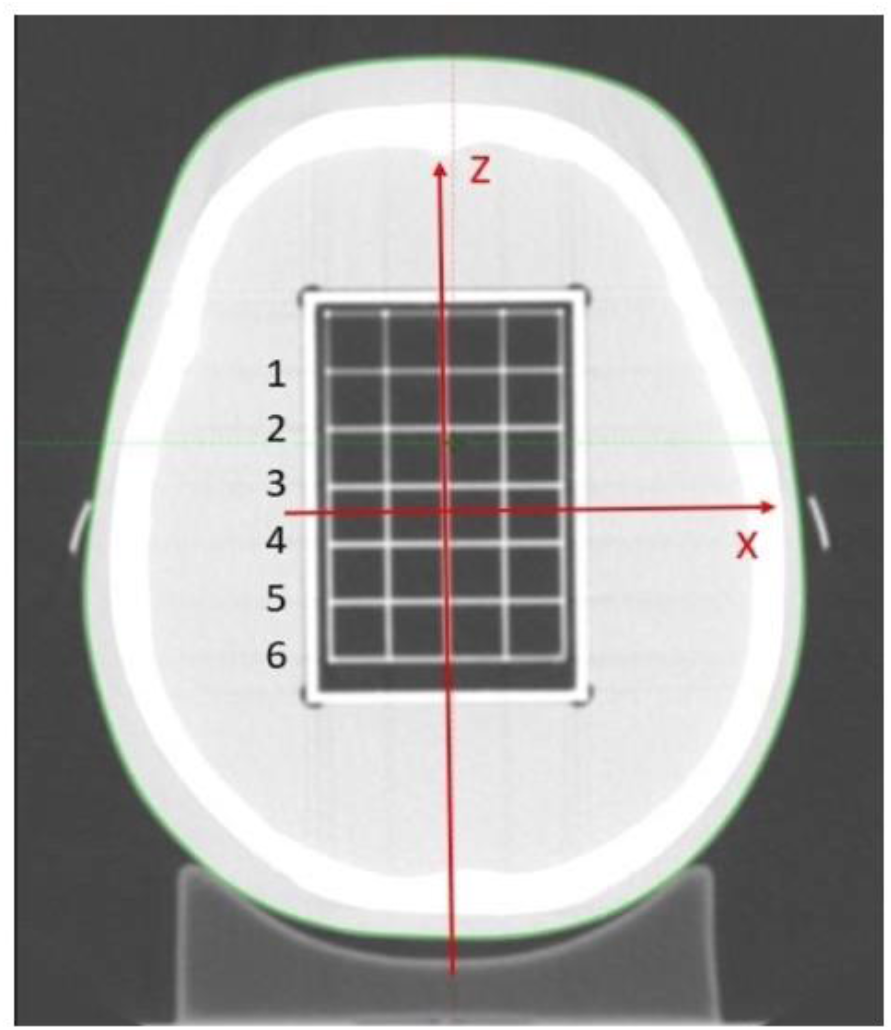
“The sequence of rows inside STEEV phantom”

## Impact on radiation dose

Simulation of the impact of geometric target shift on dose coverage yielded the following results. With a systematic positioning error corresponding to 1 mm distortion, for a hypothetical target volume of 0.1 cm^3^ (radius ∼2.9 mm), the D99% value decreased from 95% to 82% (a drop of 13 percentage points). For a large target volume of 5.2 cm^3^, the same shift caused a D99% decrease from 95% to 94.5% (less than 0.5%).

The clinical significance of distortion is inversely proportional to target volume. The calculated threshold: distortion up to and including 1 mm does not exert a significant impact on coverage for targets larger than 1.6 cm^3^. Distortion of 2 mm or greater is critical for targets smaller than 5.2 cm^3^.

## Discussion

The obtained results confirm the presence of a pronounced spatial gradient of distortion: minimal distortions at the scanner center and their increase toward the periphery. This is consistent with the physical causes of distortion – gradient field nonlinearity and edge effects described in the literature [1, 2, 4].

The distortion measurement method based on linear distances in a DICOM viewer, employed in this study, demonstrated high reproducibility. The measurement error attributable to manual delineation, amounting to ± 0.05 mm, enables reliable detection of clinically significant distortions (> 1 mm) at the periphery of the field of view. The proposed approach does not require specialized image processing software (unlike vector-based displacement methods), rendering it suitable for routine quality assurance in resource-limited clinical settings.

Validation of the custom-made ABS plastic phantom confirms its suitability as an affordable tool for routine quality control. Combined with the commercial STEEV phantom, it provides comprehensive assessment of both local and global distortions.

The obtained data are of particular significance for stereotactic radiosurgery, where targets are often small (< 2 cm^3^). As demonstrated in this study, even submillimeter distortions may lead to substantial reduction in dose coverage for such targets.

## Clinical Recommendations

- For high-precision procedures (stereotactic radiosurgery), target volumes should, whenever possible, be positioned in the central zone of the scanner (within 50 mm from isocenter), where distortion is minimal.
- For targets located at the periphery, the planning target volume (PTV) margin should be increased proportionally to the measured distortion in that region.
- For small-volume targets (< 1.6 cm^3^), distortion correction, CT verification, or patient repositioning to bring the target closer to the center is recommended.
- The distortion-versus-coordinate graphs may be utilized in clinical practice for individualized contour correction.

## Conclusion

A method for assessing geometric MRI distortion based on manual linear measurements of periodic structures in a DICOM viewer, with subsequent calculation of the difference between MRI and reference CT in Microsoft Excel, has been developed and validated. The use of two phantoms — commercial STEEV and custom-made ABS plastic — enabled distortion assessment both locally and across the entire field of view.

1. For the STEEV phantom, the maximum distortion did not exceed 0.95 mm, confirming high accuracy in the central zone.
2. For the custom-made phantom, distortions are minimal in the central zone (0.30 ± 0.05 mm), while at the periphery (> 50 mm) they reach 1.50 ± 0.05 mm or more. The maximum recorded values were 3.1 mm in the axial plane and 1.8 mm in the sagittal plane.
3. Distortion exhibits a pronounced spatial dependence: it increases with distance from the isocenter, which must be taken into account when planning irradiation of peripheral structures.
4. Distortion up to and including 1 mm is not critical for targets larger than 1.6 cm ^3^; distortion of 2 mm or greater is critical for targets smaller than 5.2 cm ^3^. For small targets, distortions may have clinically significant consequences.
5. The combination of two phantoms enables distortion assessment both locally and across the entire field of view.
6. Routine distortion control is a mandatory condition for ensuring radiotherapy quality.
7. The obtained results are being implemented in clinical practice: when planning irradiation of metastases, the planning target volume margin is adjusted according to target localization and the magnitude of distortion in that region.

## Data Availability

All data produced in the present study are available upon reasonable request to authors.

https://zenodo.org/records/20774499

## Acknowledgements

The author expresses gratitude to the supervisor, Lykova E.N., for consultation and organizational support at all stages of the work, as well as to the staff of the European Medical Centre for providing access to the equipment, scanning time, and technical support.

## Notes

### Competing Interest Statement

The authors have declared no competing interest.

